# Racial Differences in FMD

**DOI:** 10.1101/2023.02.10.23285630

**Authors:** Andrea Martinez, Alexis Okoh, Yi-An Ko, Bryan Wells

**Affiliations:** Emory University School of Medicine, Atlanta, GA; Emory Department of Medicine, Division of Cardiology, Atlanta, GA; Department of Biostatistics and Bioinformatics, Rollins School of Public Health, Emory University, Atlanta GA

## Abstract

**Background:** Fibromuscular dysplasia (FMD) is a non-atherosclerotic arteriopathy associated with stenosis, aneurysm, and dissection. We aimed to characterize racial differences in clinical presentation and diagnosis among patients with FMD at our institution.

**Methods:** We utilized an ambulatory FMD database to review demographics, clinical presentation, and diagnostic assessments of patients diagnosed with FMD within a university-affiliated healthcare system. Patients were classified as White or Non-White based on self-identified race. We evaluated race-specific differences in diagnosis and disease manifestations.

**Results:** A total of 208 patients (White: n=160 (77%); Non-White; n=48 (23%)) were included in the analysis. The time from initial FMD symptom to diagnosis was longer in Non-Whites than Whites (5.5 vs. 1.5 yrs; p<0.001), yet time from diagnosis to Emory specialist center visit was longer in Whites (3.2 vs. 1.2 yrs; p=0.035). Whites were more likely to undergo ≥ 5 multi-imaging diagnostic assessments than Non-Whites (89% vs. 73%; p=0.002). History of hypertension, stroke, & chronic kidney disease were more common in Non-Whites. FMD involvement of the internal carotid artery and upper extremity vessels were more common in Non-Whites, while Whites had more renal artery involvement.

**Conclusion:** We found racial differences in the diagnosis of FMD. The time from symptom onset to diagnosis was longer in Non-White than White FMD patients. Multimodality diagnostic imaging was more often utilized in Whites than Non-Whites. Research is needed to investigate these racial differences.

## Introduction

Fibromuscular dysplasia (FMD) is a non-atherosclerotic, non-inflammatory vasculopathy involving primarily medium-sized arteries and associated with vessel tortuosity, stenosis, aneurysm, and dissection. FMD is associated with the abnormal development of vessel media and is predominantly seen in middle aged Caucasian females (Rana, 2021). There is a concern that minorities and patients with lower socioeconomic status and limited access to healthcare are underdiagnosed and underrepresented registries and other studies. Currently, the US FMD registry shows a predominance of Caucasians being affected by the disease, with the demographics being 95.4% white, 2.2% black, 1.5% Hispanic, 0.5% Asian, and 0.5% other (Olin, 2012). However, there is no indication that FMD is a genetically white disease. On the contrary, there has been one retrospective study on cervical arterial fibromuscular dysplasia on the US–Mexican Border that suggests that FMD affecting the carotid and vertebral arteries has a similar demographic pattern across ethnicities in the United States (Qureshi, 2017). Additionally, the socioeconomic and ethnic factors that influence the disparities in access to quality healthcare, such as cost, education, and access, are well described (Riley, 2012).

The diagnosis of FMD remains a challenge since patients may be asymptomatic for a number of years. Even if symptomatic, there are a wide range of symptoms that can indicate FMD based on the vascular bed involved. Clinical manifestations of FMD may depend on the severity the pathology in an associated anatomic location. If symptoms and manifestations are present, they include migraines, hypertension, dizziness, pulsatile tinnitus, acute coronary syndromes, transient ischemic attacks, strokes, aneurysms, dissections, among others (Shah, 2021). Even though FMD is associated with these life-threatening complications, this disease is often underdiagnosed and overlooked (Rana, 2021). Early and accurate diagnosis are essential to reduce cardiovascular events associated with FMD (Shah, 2021). The mean time from first clinical symptom or sign to diagnosis of FMD is relatively long at years (Olin, 2012). FMD initial assessment and diagnosis includes evaluation of symptoms, physical examination, comprehensive imaging, and referral, all of which are directly correlated to health care affordability, geography, transportation, health literacy, access to providers with knowledge about FMD, and provider bias (Potosky, 1998; Ferguson, 1997). Thus, patients from disparate ethnic and socioeconomic backgrounds may not have the ability to receive such evaluation; and if they do, they have a higher chance of receiving an incomplete workup and therefore have a decreased rate of diagnosis of FMD and other associated conditions. Being a member of a minority group has been shown to be associated with less intense and appropriate health care (Fiscella, 2000). This study investigated the racial differences and referral patterns among patients with FMD.

## Materials and Methods

Utilizing the ambulatory Emory FMD provider database, racial differences were studied by performing a retrospective review, comparing medical data between White vs. Non-White FMD patients from 01/01/2000-01/01/2022. Given that the number of patients from minority races and ethnicities was too small to make a statistically significant comparison among all groups, Non-White patients were treated as one group. Demographics, diagnosis, treatment, and outcome variables were obtained. Additionally, further analysis was performed to help explain differences by comparing socioeconomic status based on zip code, referral clinic zip code to measure travel distance, insurance status, among others.

The Emory FMD database was utilized to perform a retrospective chart analysis of all FMD patients seen by the FMD program. The baseline disease characteristics and variables that were utilized in the analysis were: age at first FMD-related symptom, age at FMD diagnosis, time from initial symptom to diagnosis (in years), sex, self-identified race/ethnicity, insurance status, zip code, zip code from referring clinic, distance in miles from referring clinic to FMD center, mean household income based on zip code, type of FMD (focal or multifocal), symptoms at clinic presentation, significant family medical history, location and number of vascular beds involved, cardiovascular comorbidities and outcomes (prior MI, congestive heart failure, atrial fibrillation, history of stroke/TIA), number of diagnostic studies performed, and severity of outcomes and interventions (composite of MI, TIA/strokes, dissections, aneurysms). The exclusion criteria included patients under the age of 18 years, patients with no self-identified race/ethnicity available, and patients without confirmed FMD based on imaging.

To compare the abovementioned factors between White and Non-White patients with FMD, two-sample t-test and Wilcoxon test were used for continuous variables and chi-square test, and Fisher’s exact test were used for categorical variables, as appropriate.

Ethics committee/IRB of Emory University gave ethical approval for this work. Emory University’s Institutional Review Board (IRB) approval was obtained prior to the initiation of the retrospective chart review. Approval was received under expedited review. An expedited review procedure consists of a review of research involving human subjects by the Institutional Review Board chairperson or by one or more experienced reviewers designated by the chairperson from among members of the Institutional Review Board. The project posed minimal risk and therefore fit expedited review category. No annual review is required by the Institutional Review Board. A complete waiver of HIPAA authorization and informed consent was granted by the Emory University Institutional Review Board.

## Results

A total of 208 patients (White: n=160 (77%); Non-White; n=48 (23%)) were included (Table 1). For both Whites and Non-Whites, the majority of the patients were female (98% vs. 96% respectively, p=0.393). The mean age of patients was 62 ± 11 for White FMD patients and 59 ± 11 for Non-White FMD patients (p=0.095). There was no statistically significant difference between White and Non-White patients with FMD in terms of insurance status (p=0.209), household income (p=0.130), and distance from residence to Emory FMD Specialist Clinic (p=0.07). Insurance status was divided into no insurance, private insurance, and Medicare/Medicaid. Most FMD patients seen at the Emory FMD Clinic had private insurance (Whites 61% vs. Non-Whites 65%). The remaining patients had Medicare/Medicaid (Whites 38% vs. Non-Whites 31%) or no insurance (Whites 0.6% vs. Non-Whites 4%). Mean household income based on zip code was $69,503 ± 1,961 for White FMD patients and $63,313 ± 3,570 for Non-White FMD patients (p=0.130). Mean distance from patients’ main residence to clinic based on zip code was 66 ± 12.6 miles for White FMD patients and 24 ± 17 miles for Non-White patients (p=0.07).

**Table 1:** Demographics, medical history, presenting symptoms, vascular territories, and outcomes in FMD patients stratified by race.

|  | Whites<br>(n=160) | Non-whites<br>(n=48) | p-value |
| --- | --- | --- | --- |
| Age |  |  |  |
| Current Age | 62 ± 11 | 59 ± 11 | 0.095 |
| Symptom Onset | 54 ± 13 | 49 ± 14 | **0.047** |
| Diagnosis | 55 ± 12 | 55 ± 11 | 0.815 |
| Symptom to diagnosis [yrs.] | 1.5 ± 4.2 | 5.5 ± 7 | **<0.001** |
| Age seen at Emory-Specialist Center | 58 ± 11 | 55 ± 11 | 0.113 |
| Diagnosis to Specialist<br>Center [yrs.] | 3.2 ± 6.1 | 1.2 ± 3.9 | **0.035** |
| Sex [F] | 157 (98) | 46 (96) | 0.393 |
| Average Household<br>Income- Based on Zip<br>Code | 69503 ± 1961 | 63313 ± 3570 | 0.130 |
| Distance (miles)<br>Residence to Clinic | 66 ± 12.6 | 24 ± 17 | 0.07 |
| Insurance status |  |  |  |
| None | 1 (0.6) | 2 (4) | 0.209 |
| Private | 98 (61) | 31 (65) |  |
| Medicare/Medicaid | 61 (38) | 15 (31) |  |
| Medical History |  |  |  |
| Hypertension | 109 (68) | 40 (83) | **0.021** |
| HLD | 53 (33) | 13 (27) | 0.426 |
| DM | 1 (0.6) | 2 (4) | 0.106 |
| CAD | 9 (6) | 1 (2) | 0.413 |
| Tobacco use | 20 (13) | 4 (8) | 0.441 |
| M.I. | 6 (4) | 0 (0) | 0.154 |
| CVA | 13 (8) | 10 (21) | **0.021** |
| TIA | 20 (13) | 6 (13) | 1.00 |
| Dissection | 40 (25) | 3 (6) | **0.026** |
| CKD | 1 (0.63) | 12 (25) | **0.021** |
| Aneurysm | 25 (16) | 6 (13) | 0.577 |
| Family History |  |  |  |
| Aneurysm | 20 (12.5) | 5 (10) | 0.693 |
| CVA | 35 (22) | 9 (19) | 0.639 |
| FMD | 5 (3) | 0 (0) | 0.102 |
| MI | 34 (21) | 9 (19) | 0.705 |
| Symptoms |  |  |  |
| Headaches | 118 (74) | 118 (74) | 0.441 |
| Tinnitus | 88 (55) | 29 (60) | 0.506 |
| Syncope | 23 (15) | 8 (17) | 0.711 |
| Chest pain | 38 (24) | 18 (38) | 0.069 |
| Dizziness | 61 (38) | 25 (52) | 0.093 |
| Asymptomatic | 16 (10) | 2 (4) | 0.172 |
| Vessels involved |  |  |  |
| ICA | 125 (78) | 45 (94) | **0.007** |
| Vertebral | 24 (15) | 5 (10) | 0.408 |
| ECA | 1 (0.6) | 0 (0) | 0.468 |
| Coronary | 1 (0.6) | 0 (0) | 0.468 |
| Mesenteric | 11 (7) | 3 (6) | 0.818 |
| Renal | 80 (50) | 12 (25) | **0.002** |
| Upper Extremity | 0 (0) | 2 (4) | **0.015** |
| Lower Extremity | 8 (5) | 0 (0) | **0.038** |
| Diagnostic imaging |  |  |  |
| CTA Head & Neck | 146 (91) | 37 (77) | **0.013** |
| CTA Chest/Abdomen | 35 (22) | 9 (19) | 0.321 |
| CTA Abdomen | 17 (11) | 3 (6) | 0.421 |
| CTA Abdomen/Pelvis | 113 (71) | 29 (60) | **0.021** |
| US Carotid | 144 (90) | 42 (88) | 0.627 |
| US Renal | 110 (69) | 23 (48) | **0.009** |
| MRA | 103 (64) | 24 (50) | 0.076 |
| ≥ 5 Diagnostic Studies | 142 (89) | 35 (73) | **0.002** |
| Post diagnosis |  |  |  |
| MI | 40 (25) | 16 (33) | 0.261 |
| Dissection | 45 (28) | 16 (33) | 0.491 |

HLD- hyperlipidemia, DM- diabetes mellitus, CAD- coronary artery disease, MI- myocardial infarction, CVA- cerebrovascular accident, TIA- transient ischemic attack, CKD- chronic kidney disease, FMD- fibromuscular dysplasia, ICA- internal carotid artery, ECA- external carotid artery, CTA- computed tomography angiography, US- ultrasound, MRA- magnetic resonance angiography

There were comorbidities that had statistically significant differences between White and Non-White FMD patients. 68% of White FMD patients carry a diagnosis of hypertension vs. 83% of Non-White FMD patients (p=0.021). Cerebral vascular accidents (CVAs) were different between Whites and Non-Whites (p=0.021), with 8% of Whites having suffered a CVA and 21% of Non-Whites having suffered a CVA. Additionally, 25% of White FMD patients had suffered from vascular dissections vs. 6% of Non-White FMD patients, (p=0.026). 25% of Non-Whites suffered from chronic kidney disease (CKD) vs. 0.63% of Whites, (p=0.021). The other comorbidities studied, such as hyperlipidemia (p=0.426), diabetes mellitus (p=0.106), coronary artery disease (p=0.413), tobacco use (0.441), myocardial infarction (MI) (p=0.154), transient ischemic attacks (TIA) (p=1.00), and aneurysms (p=0.577) did not demonstrate significant differences between White and Non-White FMD patients. When comparing family history between White and Non-White FMD patients, there were no significant differences. The specific family histories that were studied and compared were those of aneurysms (p=0.693), CVAs (p=0.693), FMD (p=0.102), and MIs (0.705).

FMD symptoms (or lack thereof) were compared between the White and Non-White FMD patients. The difference in the mean age of first FMD symptom onset between White and Non-White FMD patients was significant, with White FMD patients having the first symptom onset at 54 ± 13 vs. Non-Whites at 49 ± 14 (p=0.047). The mean age at which patients were diagnosed with FMD is 55 ± 12 for White FMD patients vs. 55 ± 11 for Non-White FMD patients (p=0.815). The time, in years, from initial FMD symptom to FMD diagnosis was significantly different between Whites and Non-Whites (1.5 ± 4.2 vs. 5.5 ± 7 respectively, p<0.001). Additionally, the time, in years, from diagnosis to first visit to the Emory FMD Specialist Center shows a significant difference between Whites and Non-Whites (3.2 ± 6.1 vs. 1.2 ± 3.9 respectively, p=0.035). There were no statistically significant differences found between Whites and Non-Whites from the symptoms studied. This includes headaches (p=0.441), tinnitus (p=0.506), syncope (0.711), chest pain (p=0.069), dizziness (p=0.093), and no symptoms/asymptomatic (p=0.172). However, there was a difference in vessel involvement in Whites vs. Non-Whites. Internal carotid artery (ICA) involvement was present in 94% of Non-White FMD patients vs. 78% of White FMD patients, (p=0.07). Renal artery involvement also demonstrated 50% of Whites having renal artery involvement vs. 25% of Non-Whites, (p=0.002). Upper extremity arterial involvement was seen in 4% of Non-Whites vs. 0% of Whites (p=0.015), and lower extremity arterial involvement was seen in 5% of Whites vs. 0% of Non-Whites, (p=0.038). Other arterial including vertebral arteries (p=0.408), external carotid arteries (p=0.468), coronary arteries (p=0.468), and mesenteric arteries (p=0.818), did not demonstrate any significant difference between Whites vs. Non-Whites.

The types as well as the number of diagnostic imaging performed on White FMD patients vs. Non-White FMD patients was different. Overall, Whites were more likely to undergo ≥ 5 multi-imaging diagnostic assessments than Non-Whites (89% vs. 73%; p=0.002). Computed tomography angiography (CTA) of the head and neck was performed on 91% of Whites vs. 77% of Non-Whites, (p=0.013). CTA of the abdomen and pelvis was performed on 71% of Whites vs. 60% of Non-Whites (p= 0.021). Similarly, ultrasound of the renal arteries (US Renal) was performed in 69% of Whites vs. 48% of Non-Whites, (p=0.009). There was no significant difference found in the percentage of Whites vs. Non-Whites that received an ultrasound of the carotid arteries (US Carotid) (p=0.627) and magnetic resonance angiography (MRA) (p=0.076).

Lastly, there seemed to be no significant difference between Whites and Non-Whites when studying the severity of outcomes and vascular events post-FMD diagnosis. 25% of Whites suffered from a MI post-diagnosis vs. 33% of Non-Whites, (p=0.261). Additionally, 28% of Whites experienced a vascular dissection post-diagnosis, while 33% of Non-Whites experienced a vascular dissection post-diagnosis, (p=0.491).

## Discussion

This study demonstrated significant differences that require further discussion and research. When comparing the results obtained in this research to those obtained from United States’ FMD registry, there were some overarching similarities yet some differences. The mean age for diagnosis in this study is 55 years which is within the same range as that of the US FMD registry (51.9 years) (Olin, 2012). This is not an unexpected finding since FMD is known to be a disease of middle-aged females. In this study, 77% of the patients were White and 23% of the patients were Non-White. However, when comparing it to the US National FMD registry study by Olin, 95.4% of the patients are White and 4.6% are Non-White (Olin, 2012). This supports our theory that minorities are underrepresented registries and other studies. FMD patients seen at the Emory FMD specialist center are within or in close proximity to Atlanta. Atlanta has a Non-White population of 59% and Caucasian population of 41.0% as of the 2020 U.S. census, which is vastly different from the U.S. population of 75.8% Caucasians and 24.2% Non-Whites (US Census, 2020). The difference from the number of Non-Whites in the National FMD registry to the number of Non-Whites in the U.S. Census (4.6% vs. 24.2%, respectively), demonstrates that Non-White patients are likely underdiagnosed and/or are less commonly referred to academic specialty centers. Lastly, Non-White FMD patients in this Atlanta-based study are 23%, when compared to the 4.6% of the national registry, also implies the presence of geographic racial differences. Work is needed to fully analyze such differences, as well as to increase minority patient representation in national registries by referral to academic centers.

Another important finding was the difference between Whites and Non-Whites from time of initial FMD symptom to diagnosis. The mean time from first FMD symptom to FMD diagnosis on the 2012 study of the US National FMD registry is 4.7 years (Olin, 2012). The mean time found in this study was 1.5 years for Whites and 5.5 years for Non-Whites. Such a difference in time from symptom to diagnosis between Whites vs. Non-Whites is likely multifactorial, and the specific reasons are not fully understood. Further research is required to address these differences. Some potential reasons for the delay in diagnosis could be that FMD was not considered in the differential diagnosis due to under recognition of the disorder, FMD having non-specific symptoms that can be caused by other more common disorders (such as primary hypertension, tension headache, etc.), and increased obstacles within the social and healthcare system for Non-Whites compared with Whites. Regardless of the specific reasons leading to such a discrepancy, it is likely that such a delay in time to diagnosis in Non-Whites has potential negative repercussions in the patients’ care and outcomes (Olin, 2008; Kim, 2008).

Additionally, there was a difference in time from FMD diagnosis to first Emory FMD Specialist visit, with Whites having a longer mean time than Non-Whites. One of the potential reasons behind this difference is that these variables look at the time difference between diagnosis to the Emory Specialist Center specifically, not just any FMD specialist center. Some individuals, many of which are White, visit the Emory FMD Specialist Center for a second opinion or further/transitioning medical care, therefore not accurately representing the time from diagnosis to any FMD specialist center. This theory is further supported by the trend of White FMD patients traveling longer distances to the Emory FMD center compared to Non-White patients. Although it was shown to have a statistically significant differences, the difference in miles traveled does demonstrate a trend that White patients travel further. Furthermore, Whites having a longer mean time from FMD diagnosis to FMD Emory Specialist visit can be potentially further explained by the fact that patients may be seeing another vascular specialist within Emory who is not affiliated with the Emory FMD Program (ie, vascular surgery or neurology).

Medical history and comorbidities also showed some differences. More Non-Whites suffered from hypertension, CVAs, and CKD compared to Whites. Although, this is not a surprising finding since, in the general U.S. population, blood pressure control rates are lower in Black, Hispanic, and Asian adults as compared to White adults, and Black adults have higher hypertension prevalence (Aggarwal, 2021). Similarly, Black adults have a higher incidence of strokes compared to Whites, and a higher incidence of CKD and faster progression of CKD to end-stage kidney disease compared to White adults (Kleindorfer, 2010; Chu, 2021). The higher presence of those comorbidities in Non-Whites could also potentially be due to the delay in diagnosis, and therefore delay in treatment since FMD can cause and present in such a manner. Further emphasizing why timely diagnosis and treatment is important for FMD. Additionally, it is not yet understood why dissections were significantly higher in Whites vs. Non-Whites. A previous abstract found similar findings, with black FMD patients being less likely than white FMD patients to have arterial dissections but had a higher percentage of aortic dissections (Shah, 2021). It is known that dissections and aneurysms occur more frequently in FMD patients compared to their counterparts in the general population (Olin, 2012), but there is no literature that explain why dissections are more common in White FMD patients. A potential explanation for this discrepancy is that Non-Whites have less frequent, and fewer imaging performed, leading to an underdiagnosis of dissections. Further research is required to further understand the differences in comorbidities and outcomes in FMD patients.

There were also some significant differences in the diagnostic testing performed in Whites vs. Non-Whites, as well as vessel involvement. White patients were more likely to undergo CTAs of the head and neck, CTAs of the abdomen and pelvis, ultrasounds of the renal arteries, and were more likely to receive five or more diagnostic studies than Non-Whites. In terms of vessel involvement, Whites have a significantly higher involvement of the renal and lower extremity vessels, which can explain the higher number of renal ultrasounds performed in Whites vs. Non-Whites. On the other hand, Non-Whites have a higher involvement of ICAs and upper extremity vessels, which is inconsistent with CTAs of the head and neck being performed more frequently in Whites. The significance and reasoning of such differences has not been fully investigated, and therefore is not well understood. A potential explanation to explain such a difference is that Whites receive a more thorough imaging screen (“brain to pelvis”) to evaluate all the potential vessels involved in FMD, therefore receiving several diagnostic imaging studies, while Non-Whites receive diagnostic imaging in the anatomical location of the current symptoms or condition. With Non-White patients receiving less imaging studies, they are likely underdiagnosed compared with White patients.

FMD is not the only disease where such racial differences are seen. This is a phenomenon that is not unfamiliar to medicine and has become the subject of study for the past few years. For instance, spontaneous coronary artery disease (SCAD) has similarly been shown to have a low number of Non-White patients in the iSCAD national registry (Wells, 2022). Additionally, that same study found significant differences in White vs Non-White patients in terms of manifestations (STEMI vs. NSTEMI), time to hospital presentation, and time to angiography being performed (Wells, 2022). Similarly, peripheral artery disease (PAD) has also been proven to have significant racial differences. Black Americans were shown to be disproportionally affected by PAD, while being less likely to be timely diagnosed and receive a promptly treatment (Hackler, 2021). Additionally, other studies further expanded on such racial disparities by determining that Black and Hispanic PAD patients are more likely undergo amputations compared to their White counterparts (Rowe, 2010). These studies further emphasize the racial differences present in medicine, with an overarching theme of minorities and underserved communities being less likely to receive adequate care.

This study has limitations that need to be considered. The number of individuals in the Emory FMD registry is low enough where Non-Whites had to be grouped together, therefore not allowing there to be a comparison between each race and ethnicity. Additionally, there may be a referral bias in the sense that the most symptomatic FMD patients and patients with previous vascular events are more likely to be evaluated at the FMD Specialist Center, making it difficult to determine if this data represents undiagnosed or less symptomatic individuals. As the Emory FMD database matures and more providers are knowledgeable about FMD, the data would be more representative and accurate. Furthermore, patients under the age of 18 years are excluded from this study since pediatric FMD is unique and symptomatically diverges from adult FMD, making this data not applicable for pediatric FMD. Additional limitations include the fact that we are analyzing data from a single center, which could potentially show data that is not generalizable to the entire U.S. FMD population. Even though certain confounding biases were considered, such as analyzing patients’ zip codes, distance traveled, insurance status, and income, there are always other risk factors that may be present that were not measured. More specifically, since there was no adjusting for multiple testing (prone to inflated type I error since many tests were performed), and only univariate analysis was performed, it justifies this as an exploratory study unable to account for confounding. Finally, this study reveals only associations and not causation, therefore the reasoning behind these differences is not fully understood, and further research is needed.

## Conclusion

This study found significant racial differences in the time to diagnosis, comorbidities, manifestations, and screening among patients with FMD. The time from symptom onset to diagnosis was longer in Non-White FMD patients when compared to White FMD patients. Multimodality diagnostic imaging was more often utilized in Whites than Non-Whites. More Non-Whites suffered from hypertension, CVAs, and CKD compared to Whites. Additionally, the difference in population of the FMD National database compared to the patients seen at the Emory FMD Specialist Clinic, with the Emory Clinic having a much larger number of Non-White patients, suggests geographic differences of FMD and the need for more representation of underrepresented minorities in registries and other studies. Research is needed to further investigate these racial differences.

## Data Availability

The data that support the findings of this study are available from the corresponding author, AM, upon reasonable request.

## Author Contributions

A.M: created, designed, and obtained IRB approval for the project. Performed the retrospective chart review. Wrote manuscript.

B.W.: guided IRB approval process. Created and built the Emory FMD provider database. Reviewed manuscript.

Y.K: provided statistical guidance.

A.O. provided statistical guidance and performed the statistical analysis.

---

**Figure 1.**
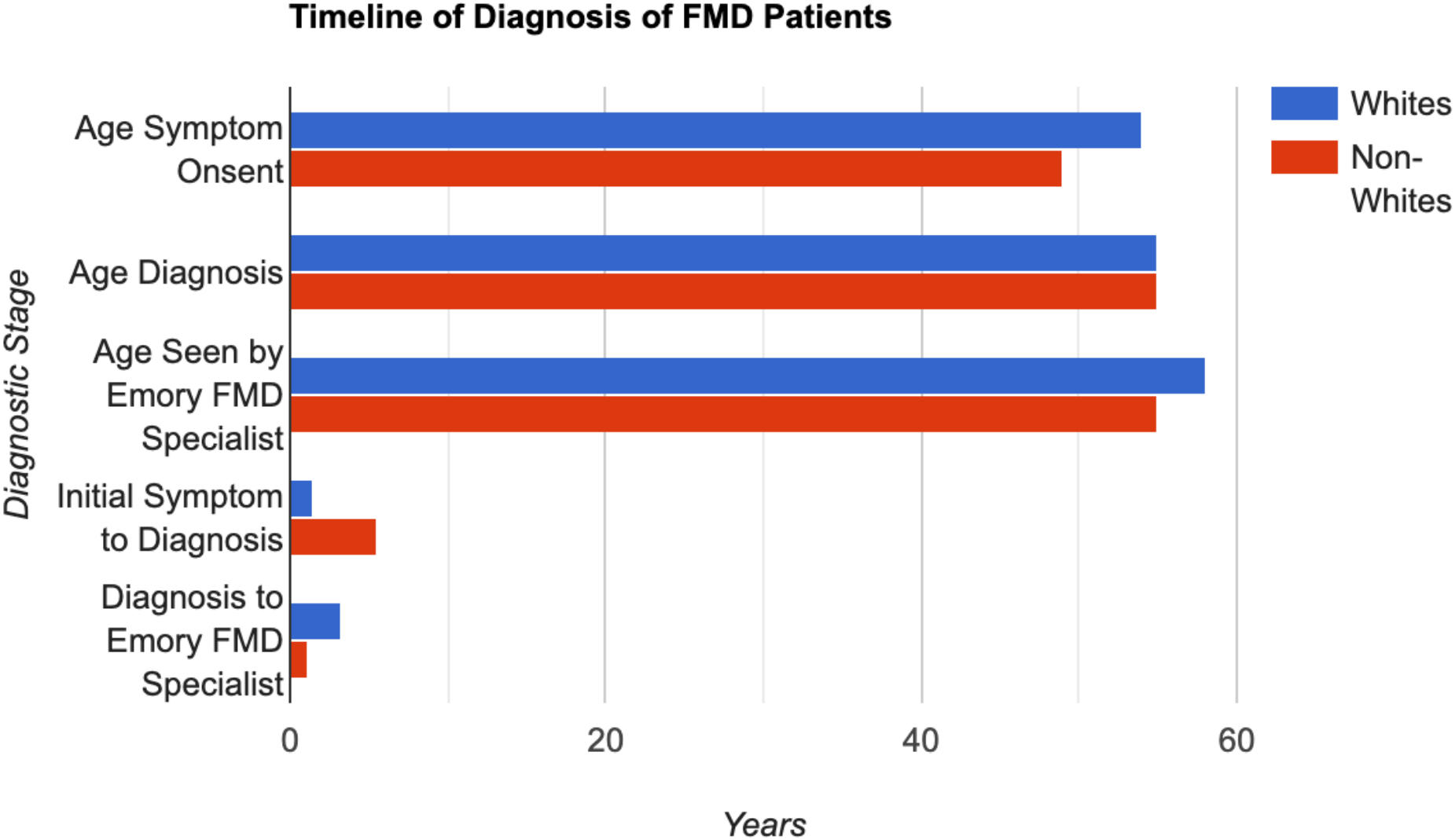
Timeline of Diagnosis of FMD Patients, Whites vs. Non-Whites.

**Figure 2.**
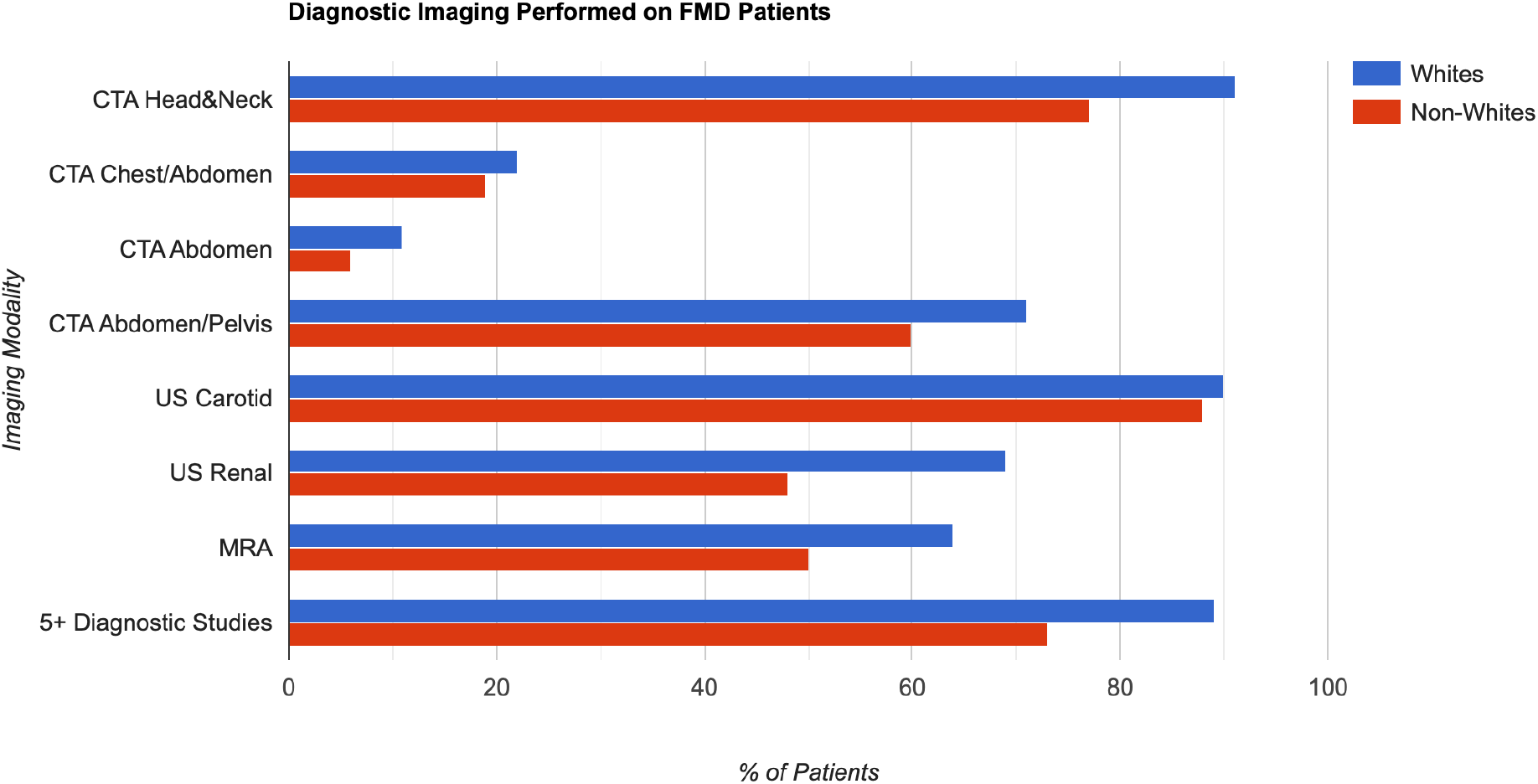
Diagnostic Imaging Performed on FMD Patients, Whites vs. Non-Whites. CTA- computed tomography angiography, US- ultrasound, MRA- magnetic resonance angiography

